# SARS-CoV-2 asymptomatic and symptomatic patients and risk for transfusion transmission

**DOI:** 10.1101/2020.03.29.20039529

**Authors:** Victor M. Corman, Holger F. Rabenau, Ortwin Adams, Doris Oberle, Markus B. Funk, Brigitte Keller-Stanislawski, Jörg Timm, Christian Drosten, Sandra Ciesek

## Abstract

Oral swabs, sputum and blood samples from 18 patients with SARS-CoV-2 infection were examined using real-time reverse transcription polymerase chain reaction (RT-PCR) testing. Whereas oral swabs or sputum from the lower respiratory tract were tested RT-PCR positive in all patients, RNAemia was neither detected in 3 patients without symptoms nor in 14 patients with flu-like symptoms, fever or pneumonia. The only patient with RNAemia suffered from acute respiratory distress syndrome (ARDS) and was artificially ventilated in an intensive care unit. Risk for SARS-CoV-2 transmission through blood components in asymptomatic SARS-CoV-2 infected individuals therefore seems negligible but further studies are needed.

## Introduction

As of March 2020, the SARS-CoV-2 pandemic that originated in Wuhan, Hubei Province, China, is quickly spreading in many countries of the world.

Huang et al. ^1^ reported RNAemia in 6 of 41 symptomatic Wuhan patients (15%) with laboratory-confirmed SARS-CoV2 infection. Zhang et al. ^2^ showed for 15 Wuhan patients diagnosed with viral pneumonia caused by SARS-CoV-2 that shedding may occur through multiple routes (respiratory, fecal–oral or body fluids); six out of the 15 patients (40%) were blood positives. In addition, Kim et al. ^3^ reported on intermittently positive blood RT-PCR (real-time reverse transcription polymerase chain reaction) results in 2 SARS-CoV-2 patients in South Korea with mild and moderate symptoms.

In Germany, blood donors have to be at least 18 years old and should not be older than 65 years of age. In addition, volunteers will not be admitted to blood donation if they show signs and symptoms of an infection (e.g. fever, flu-like symptoms). With these requirements and the additional donor screening, a high national safety standard with regard to the prevention of transfusion-related viral infections has been achieved in recent years ^4^.

However, with respect to the safety of blood donation, the questions arise whether Corona virus disease 2019 (Covid-19) can be transmitted through blood and how long SARS-CoV-2 positive patients with minimal symptoms have to be deferred from blood donation.

## Patients and methods

We report on molecular detection of SARS-CoV-2 in 18 patients 2 of whom were infected in China and evacuated to Germany on February 1 ^5^. In 16 patients, the transmission in Germany is most likely^6^. Information on the patient population was published elsewhere^5,6^ and SARS-CoV-2 testing targeting the E and RNA-dependent RNA polymerase gene was performed as previously described by Corman et al. ^7^. In patients with minor symptoms, laboratory tests were carried out during the two-week quarantine period, in those with severe symptoms, tests were done during inpatient treatment.

## Results

Twelve of the 18 patients were males and six females. Three patients were asymptomatic, six presented with flu-like symptoms, five had flu-like symptoms as well as fever, two suffered from pneumonia, and one patient needed artificial respiration because of acute respiratory distress syndrome (ARDS) (Table 1). Three of the 18 patients fulfilled the requirements for blood donation in Germany. Whereas oral swabs or sputum from the lower respiratory tract were tested RT-PCR positive in all patients, RNAemia was only detected in one out of seven serum/plasma samples taken from the patient with ARDS.

**Table 1.**
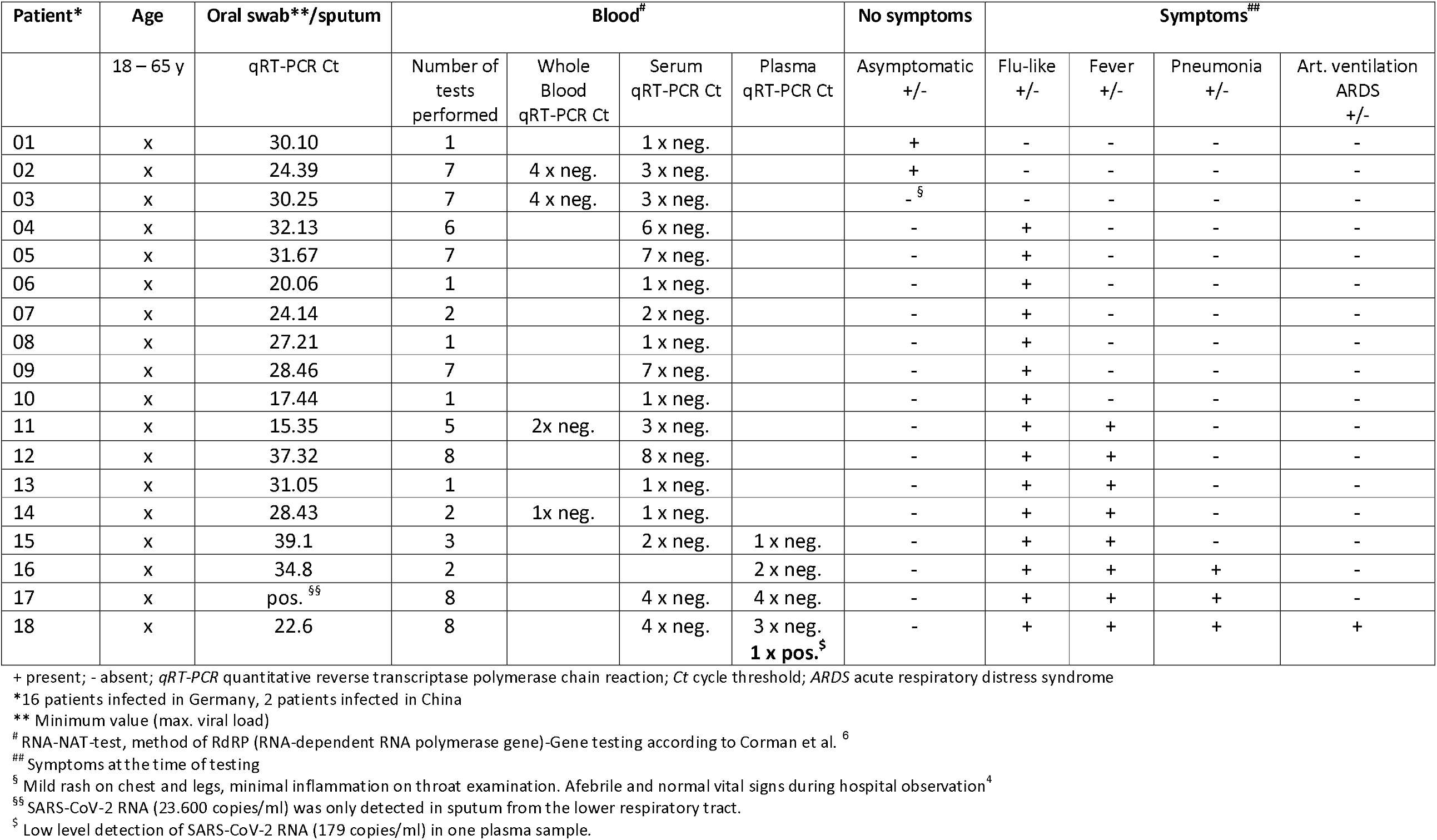
Molecular detection of SARS-CoV-2 in 18 patients with asymptomatic or symptomatic infection

## Discussion

We presented data on 18 SARS-CoV-2 infected patients, 3 patients without specific symptoms and 15 with symptoms of different severity. SARS-CoV-2 genomes were only detected in one of 77 blood samples examined. The patient with RNAemia suffered from ARDS and was artificially ventilated in an intensive care unit. In asymptomatic patients who are eligible for blood donation as well as patients with flu-like symptoms and fever, no SARS-CoV-2 RNA could be detected in the blood or serum despite a clearly positive result in all throat swabs.

Our findings are in line with published data ^8^ and confirm that SARS-CoV-2 infection may go without noticeable manifestation of clinical symptoms. Furthermore, Chen et al. ^9^ detected SARS-CoV-2 genome in the blood of 6 out of 57 Chinese patients and found RNAemia to be associated with a more severe clinical picture. In addition, detectable serum SARS-CoV-2 RNAemia was found to be closely linked to an elevated interleukin 6 (IL-6) level in critically ill Covid-19 patients^10^.

Since there is currently no recognized external standardization of the SARS-CoV-2 RT-PCR methods, a direct comparison of the results of Chinese and German studies is not possible. Of note, RNAemia is not equivalent to infectiousness, i.e. despite the presence of RNA it is unknown whether blood can harbor intact viruses that are able to infect tissues and, when transfused, can cause hematogenous transmission.

Hitherto, similarly to the other two coronaviruses that have emerged over the past 20 years (SARS, the Severe Acute Respiratory Syndrome Coronavirus ^11,12^ and MERS-CoV, causing Middle East Respiratory Syndrome^13^, no hematogenous transmissions have been documented for SARS-CoV-2. Due to a lack of evidence indicating a risk for transfusion transmission of SARS-CoV-2, the American Association of Blood Banks (AABB), the Food and Drug Association (FDA), and the Centers for Disease Control and Prevention (CDC) are not recommending any action by blood collection establishments at this time (March 3, 2020)^14^. Our data support this recommendation.

## Conclusion

Among 18 patients with RT-PCR confirmed SARS-CoV-2 infection included in our study only one critically ill individual had RNAemia. Based on these limited data, there is no measurable risk for SARS-CoV-2 transmission through blood components in asymptomatic SARS-CoV-2 infected individuals. As no cases of transmission due to transfusion of blood products have been reported for SARS, MERS-CoV and SARS-CoV-2, and considering that patients with symptoms of infectious disease will not be admitted to blood donation in Germany, the risk for transfusion transmission of SARS-CoV-2 is supposed to be negligible. However, well-powered studies are needed to further evaluate the potential risk of hematogenous transmission and to determine the appropriate referral time for blood donation of recovered SARS-CoV-2 patients.

## Data Availability

All essential data were obtained in the participating institutes for virology and are shown in the text and table.

## Competing Interest Statement

The authors have declared no competing interest.

## Ethical approval statement

All patients provided informed consent to the use of their data and clinical samples for the purposes of the present study. Institutional review board clearance for the scientific use of patient data has been granted to the treating institution by the Ethikkommission der Medizinischen Fakultät der Ludwig Maximillians Universtität München and by the University Hospital’s ethics committee, Johann Wolfgang Goethe Universtität Frankfurt/Germany.

## Funding Statement

This work was funded by the Paul-Ehrlich-Institut (Competent authority/ Germany).

### Acknowledgements

We thank Susanne Müller, Gabriele Ruppert-Seipp and Philipp Berg for technical assistance.

